# Ceramides as risk markers for future cardiovascular events and all-cause mortality in long-standing type 1 diabetes

**DOI:** 10.1101/2022.12.09.22283278

**Authors:** Asger Wretlind, Viktor R. Curovic, Tommi Suvitaival, Simone Theilade, Nete Tofte, Signe A. Winther, Tina Vilsbøll, Henrik Vestergaard, Peter Rossing, Cristina Legido-Quigley

**Author notes:** Corresponding author: Cristina Legido-Quigley, Steno Diabetes Center Copenhagen and King’s College London, Phone nr.

## Abstract

**Background:** Ceramides are lipid molecules involved in inflammation-related signaling. Recent studies have shown that higher amounts of specific circulating ceramides and ceramide ratios are associated with future development of cardiovascular (CV) disease (CVD). In persons with longstanding type 1 diabetes (T1D), we investigated if serum ceramide levels and ratios predicted CVD, kidney failure and all-cause mortality.

**Methods:** We included 662 participants with T1D from 2009-2011. Health registry data was obtained at a 6-year follow-up. Baseline serum samples were analyzed using liquid chromatography-mass spectrometry. Six predefined ceramide levels were measured and ratios calculated. Adjusted Cox regression analyses were carried out to investigate ceramide levels in relation to future CV events (CVE), kidney failure and all-cause mortality.

**Results:** Ceramide ratio cer(d18:1/18:0)/cer(d18:1/24:0) was significantly associated with risk of CVE (HR = 1.33, P = 0.01) and all-cause mortality (HR = 1,48, P = 0.01) before and after adjustments. All five investigated ceramide ratios were associated with kidney failure, before adjusting for the kidney markers estimated glomerular filtration rate and urinary albumin excretion rate.

**Conclusions:** Specific ceramides and ratios associated with 6-year cardiovascular risk and all-cause mortality in a T1D cohort. The ratio of cer(d18:1/18:0)/cer(d18:1/24:0) was an especially robust indicator. This highlights the strength of ceramide association with vascular complications and presents a new potential tool for early risk assessment if validated in other cohorts.

**Trial Registration:** ClinicalTrial.gov identifier: NCT01171248

## Introduction

The number of people living with type 1 diabetes (T1D) is rising and estimates foresee further increase in the coming years (1). Two major complications to T1D are cardiovascular (CV) disease (CVD) and chronic kidney disease (CKD), the former being the leading cause of death in individuals with T1D (2,3). Individuals with T1D and diabetic kidney disease are subjected to an increased mortality risk (4). Diabetes, CVD and kidney disease are closely interlinked sharing risk factors and molecular mechanisms (5,6).

Identification of biomarkers which stratifies those at highest risk for these complications would allow early intervention with optimization of risk factors and potentially organ protective therapies, as has been seen in type 2 diabetes (T2D) with non-steroidal mineralocorticoid receptor antagonists, glucagon-like peptide 1 receptor agonists (GLP-1RA) and sodium glucose cotransporter 2 inhibitors (SGLT2i)(7,8).

Recent studies have found a small number of specific ceramides associated with CV outcomes and mortality years before onset of the clinical disease. Specifically, Cer(d18:1/16:0), Cer(d18:1/18:0), Cer(d18:1/24:0) and Cer(d18:1/24:1), here referred to as cer16, cer18, cer24:0 and cer24:1 respectively, were reported as biomarkers of cardiovascular outcomes (9–11). Cer(d18:1/22:0) (cer22) have also been associated with CVD and all-cause mortality (12–14). It has been suggested that ceramides play an active role in progression of CVD, for instance through the promotion of reactive oxygen species in cardiomyocytes and endothelial cells (15).

The levels of these specific ceramides are also increased in individuals with T2D, and have been linked to insulin resistance (16–19) as well as to an increased risk of developing T2D (20–22). However, evidence for ceramide levels as risk-predictors for CVD in T1D cohorts is lacking. In this study we set out to evaluate these biomarkers’ ability to predict CVD, kidney failure, and all-cause mortality in a prospective study of 662 individuals with T1D.

## Research design and methods

### Participants

This study is based on a prospective cohort study of participants with T1D recruited at the Steno Diabetes Center Copenhagen outpatient clinic between 2009 and 2011, described in full by Theilade *et al*. (23). A follow-up study was carried out in 2017 obtaining information about hospitalization and death from the Danish National Heath Register and the Danish National Death Register, as well as carrying out mass spectrometry-based lipidomics analysis using serum samples collected at the start of the original study, details of the follow-up protocol have been reported by Tofte el al. (24).

The original study is registered at ClinicalTrial.gov under the identifier: NCT01171248, it holds ethical approval from the Danish National Committee on Biomedical Research Ethics (2009-056). The follow-up protocol was approved The Ethics Committee E, *Region Hovedstaden*, Denmark. Both the original study and the follow-up protocol adhere to principles of the Declaration of Helsinki. Written and informed consent was given by the participants before inclusion in the study.

### Lipid analysis

Untargeted lipidomics analysis of baseline serum was carried out using a protocol as previously described (24,25). The raw mass spectrometry data was analyzed for ceramide amounts as follows: pre-processing was carried out with MZmine 2 v.2.28 (26) (see technical details in Supplementary Material). Six ceramides of interest, namely, cer16, cer18, cer20, cer22, cer24:0 and cer24:1, were targeted using their respective water loss adduct [M-H2O+H]^+^ as the quantifier ion and the protonated adduct [M+H]^+^ as a qualifier ion with matching retention time (27). Final preprocessing was performed in R v.4.2.0 (28). Semi-quantification was achieved by comparing ceramide peak areas to the peak area of an exogenous pure standard Cer(d18:1/17:0) spiked in all samples for a final concentration of 2µg/ml. Outliers were defined as measures more than three standard deviations away from the median. R code for preprocessing can be found on Github: https://github.com/Asger-W/Profil-Ceramides.

### Statistics

All statistics and visualizations were produced in R, and the code for the statistical analysis can be found on Github. Clinical characteristics presented as n (%), mean (SD) or median [IQR] were compared between individuals developing CV events (CVE) or those who didn’t using Fisher’s exact test and compiled into a table with the tableone package. Survival analysis was performed with the survival and survminer packages. Cox regression were carried out for each ceramide and ratio to cers24:0, using three levels of adjustments: a crude model without adjustments, a model with level 1 adjustment, adjusted for age, sex, body mass index (BMI), low-density lipoprotein (LDL), triglycerides, systolic blood pressure, glycated hemoglobin A1c (HbA_1C_), history of CVD, smoking status and statin use. The model with final adjustments (level 2 adjustments) included the estimated glomerular filtration rate (eGFR) and log-transformed urinary albumin excretion rate (UAER) in addition to the confounders in the level 1 adjusted model. CVE are a composite of several outcomes, as previously defined (29), namely CV mortality, coronary artery disease including non-fatal myocardial infarction and coronary revascularization (percutaneous arterial intervention or coronary bypass grafting), non-fatal stroke, and peripheral arterial interventions including amputations. Kidney failure was defined as either receiving dialysis, kidney transplantation or having a eGFR ≤15 mL/min/1.73 m^2^ (24). Albuminuria status was defined by UAER, normoalbuminuria was defined as having a UAER below 30 mg/g, moderate increase in albuminuria as UAER between 30-299 mg/g and severe increase in albuminuria as UAER ≥300 mg/g. The variables used for adjustment were themselves modeled with Cox regression, in one model without adjustment and in one model with the same adjustments as the level 2 adjusted model except for the interrogated variable. Correlation matrix of Pearson correlations was produced and drawn as heatmaps with the ggcorrplot package.

## Results

### Baseline characteristics

Here we investigated a prospective cohort of 662 participants with T1D, baseline lipid measurements and follow-up data regarding disease and death with median follow-up time of 6.3 years. The mean (SD) age was 55 (13) years, the diabetes duration was 33 (16) years and 45 % were women. A total of 94 participants experienced a cardiovascular event, 23 progressed to kidney failure and 58 died. Baseline characteristics of the total and subpopulations with and without CVE are summarized in Table 1 and ceramide measures in Table 2. Participants experiencing a CVE had generally higher lipid levels typically associated with CV risk. Total cholesterol, LDL and very low-density lipoprotein (VLDL) were higher compared to the groups without CVE, but not significantly so, while triglycerides and cer16 and cer18 levels were significantly higher at baseline in the CVE group compared to the non-CVE group (mean (SD) = 2.32 (0.41) ng/ml vs. 2.19 (0.45) ng/ml and 1.36 (0.41) ng/mol vs. 1.24 (0.35) ng/ml respectively; p = 0.01 and 0.002, respectively).

**Table 1.** Clinical characteristics

|  | Overall | No CVE | CVE | p-test |
| --- | --- | --- | --- | --- |
| n | 662 | 568 | 94 |  |
| Age (years) | 54.6 (12.7) | 53.6 (12.9) | 60.7 (9.5) | <0.001 |
| Diabetes duration (years) | 32.7 (15.9) | 31.3 (15.8) | 41.3 (13.1) | <0.001 |
| Women | 296 (45%) | 264 (47%) | 32 (34%) | 0.038 |
| Smoking | 137 (21%) | 116 (20%) | 21 (22%) | 0.774 |
| BMI (kg/m<sup>2</sup>) | 25.4 (5.8) | 25.4 (6.0) | 25.7 (4.2) | 0.619 |
| Previous CVD | 139 (21%) | 85 (15%) | 54 (57%) | <0.001 |
| Retinopathy:<br>No /mild-moderate non-<br>proliferative / proliferative | 140 (21%) / 275 (42%)<br>/<br>120 (18%) | 133 (23%) /<br>246 (43%) /<br>98 (17 %) | 7 (7%) /<br>29 (31%) /<br>22 (23%) | <0.001 |
| UAER (mg/g) | 3.46 [2.46-6.11] | 3.19 [2.41-5.58] | 5.37 [3.68-10.38] | <0.001 |
| HbA<sub>1c</sub> (mmol/mol) | 64.3 (12.7) | 63.8 (12.6) | 67.7 (12.5) | 0.006 |
| HbA<sub>1c</sub> (%) | 8.0 (1.2) | 8.0 (1.2) | 8.3 (1.1) | 0.006 |
| Hemoglobin (mmol/L) | 8.5 (0.9) | 8.5 (0.8) | 8.2 (1.0) | 0.004 |
| Total cholesterol (mmol/L) | 4.7 (0.9) | 4.7 (0.8) | 4.8 (1.0) | 0.126 |
| HDL (mmol/L) | 1.7 (0.5) | 1.7 (0.5) | 1.6 (0.6) | 0.083 |
| LDL (mmol/L) | 2.5 (0.8) | 2.4 (0.7) | 2.6 (0.9) | 0.069 |
| VLDL (mmol/L) | 0.5 (0.3) | 0.5 (0.3) | 0.6 (0.3) | 0.017 |
| Triglyceride (mmol/L) | 1.13 (0.66) | 1.09 (0.58) | 1.35 (1.01) | <0.001 |
| eGFR (ml/min/1.73m<sup>2</sup>) | 81.53 (25.51) | 83.95 (24.69) | 67.00 (25.68) | <0.001 |
| hsCRP (mg/L) | 3.41 (7.02) | 3.36 (7.20) | 3.70 (5.83) | 0.666 |
| Systolic blood pressure (mmHg) | 132 (17) | 131 (17) | 137 (19) | 0.001 |
| Diastolic blood pressure (mmHg) | 74 (9) | 75 (9) | 73 (10) | 0.067 |
| Below albuminuria / moderately increased / severely increased (<30 / 30-299 / ≥300 mg/g) | 308 (47%) / 165 (25%) / 189 (29%) | 290 (51%) / 133 (23%) / 145 (26%) | 18 (19%) / 32 (34%) / 44 (47%) | <0.001 |
| RAAS treatment* | 445 (67%) | 358 (63%) | 87 (93%) | <0.001 |
| Antihypertensive treatment | 475 (72%) | 382 (67%) | 93 (99%) | <0.001 |
| Beta blocker treatment | 85 (13%) | 58 (10%) | 27 (29%) | <0.001 |
| Calcium channel blocker treatment | 202 (31%) | 155 (27%) | 47 (50%) | <0.001 |
| Insulin pump user | 57 (9%) | 52 (9%) | 5 (5%) | 0.303 |
| Daily insulin dose (IU) | 48.7 (35.0) | 48.8 (36.2) | 47.9 (26.3) | 0.824 |
| Statin treatment | 397 (60%) | 321 (57%) | 76 (81%) | <0.001 |
| Aspirin or clopidogrel treatment | 349 (53%) | 273 (48%) | 76 (81%) | <0.001 |
| Diuretic treatment | 334 (51%) | 260 (46%) | 74 (79%) | <0.001 |
| Thiazide treatment | 186 (28%) | 159 (28%) | 27 (29%) | 0.982 |
| Loop diuretic treatment | 149 (23%) | 105 (19%) | 44 (47%) | <0.001 |
| MRA treatment | 28 (4%) | 17 (3%) | 11 (12%) | <0.001 |

Data presented as n (%), mean (SD) or median [IQR]. Groupwise comparison between the two treatments were tested using Fisher's exact test. BMI: Body mass index, CVD: Cardiovascular disease, UAER: Urine albumin excretion, HbA<sub>1c</sub>: Hemoglobin A1c, HDL: High-density lipoprotein, LDL: Low-density lipoprotein, VLDL: Very low-density lipoprotein, eGFR: Estimated glomerular filtration rate, hsCRP: High-sensitivity C-reactive protein, RAS: Renin angiotensin system, MRA: Mineralocorticoid receptor antagonist. \*Spironolactone is given separately which blocks aldosterone.

**Table 2.** Ceramide measures

|  | Overall | Individuals without CVE | Individuals who developed CVE | p-test |
| --- | --- | --- | --- | --- |
| n | 662 | 568 | 94 |  |
| Cer16 ng/ml | 2.21 (0.44) | 2.19 (0.45) | 2.32 (0.41) | 0.01 |
| Cer18 ng/ml | 1.26 (0.36) | 1.24 (0.35) | 1.36 (0.41) | 0.002 |
| Cer20 ng/ml | 1.78 (0.66) | 1.78 (0.66) | 1.83 (0.71) | 0.496 |
| Cer22 ng/ml | 10.26 (4.28) | 10.24 (4.22) | 10.43 (4.65) | 0.686 |
| Cer24:0 ng/ml | 39.99 (16.29) | 39.94 (16.00) | 40.30 (18.06) | 0.842 |
| Cer24:1 ng/ml | 19.55 (7.44) | 19.32 (7.20) | 20.93 (8.66) | 0.052 |
| Ratio cer16/cer24:0 | 0.06 (0.02) | 0.06 (0.02) | 0.07 (0.03) | 0.102 |
| Ratio cer18/cer24:0 | 0.03 (0.01) | 0.03 (0.01) | 0.04 (0.01) | 0.009 |
| Ratio cer20/cer24:0 | 0.05 (0.01) | 0.05 (0.01) | 0.05 (0.01) | 0.218 |
| Ratio cer22/cer24:0 | 0.26 (0.04) | 0.26 (0.04) | 0.26 (0.04) | 0.437 |
| Ratio cer24:1/cer24:0 | 0.51 (0.11) | 0.50 (0.11) | 0.54 (0.12) | 0.002 |

Data is presented as mean (SD). Groupwise comparison between the two groups were tested using Fisher's exact test.

### Ceramide levels and longitudinal endpoints

Cox regression analysis was carried out on the six investigated ceramides to test their association with CVE, kidney failure and all-cause mortality (Figure 1).

**Figure 1.**
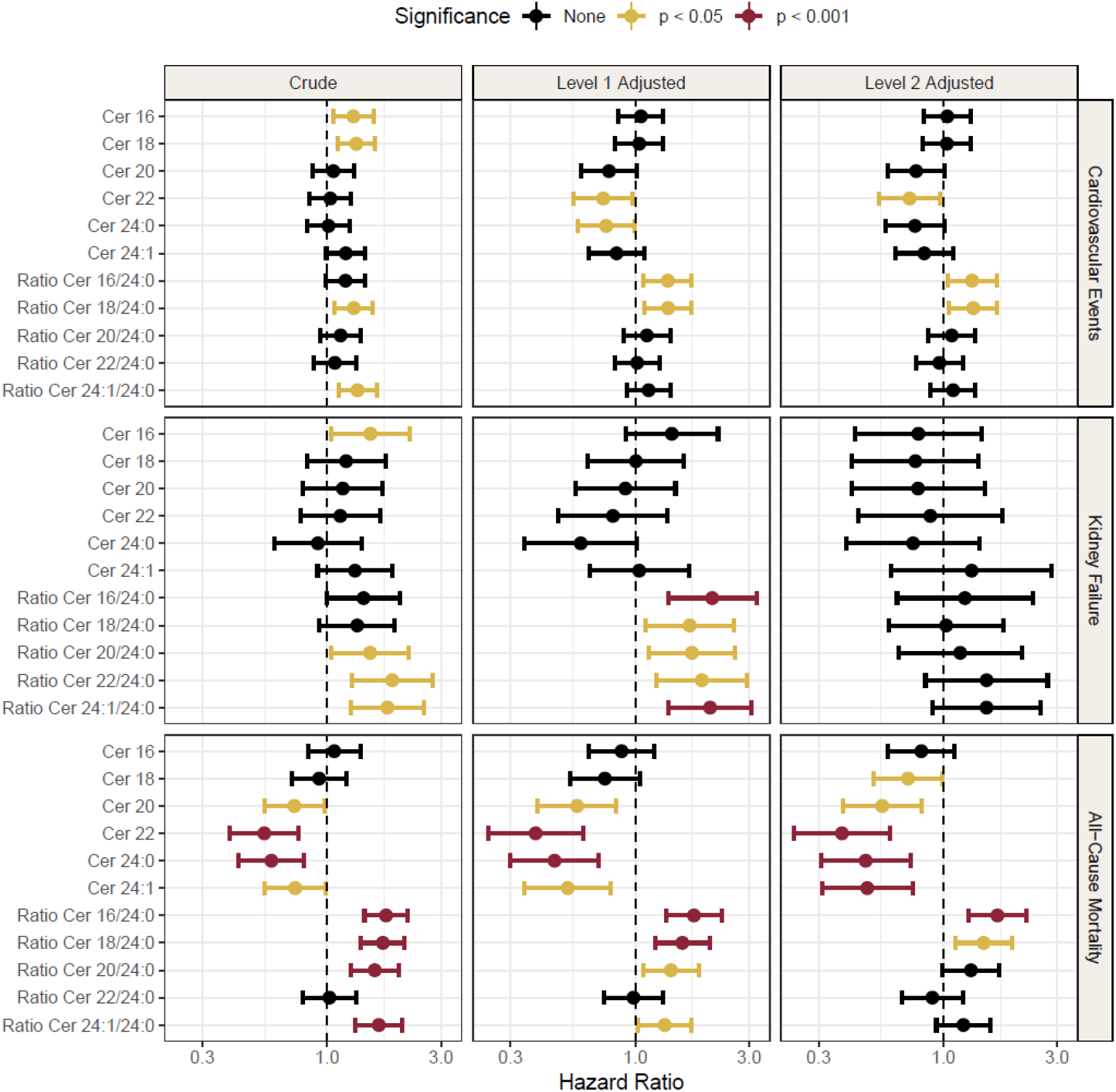
Forest plot of hazard ratios for ceramide and ratios for outcomes of cardiovascular events, kidney failure and mortality. The crude models are unadjusted, level 1 adjusted for age, sex, BMI, LDL, triglycerides, systolic blood pressure, HbA_1C_, history of CVD, smoking status and statin use. Level 2 adjusted for all the same variables as level 1, but also included eGFR and UAER. Hazard ratios (HRs) are reported per doubling of the log10 ceramide.

For cardiovascular outcomes, the ratio of cer18/cer24:0 was the only measure that was significantly associated at all levels of adjustments (level 2 adjusted model: HR = 1.33, 95% CI =1,06-1.68, P =0.01). Cer16, cer18 and the ratio cer24:1/cer24:0 were significantly associated with CVE in crude models, but not in adjusted models. Cer22 (HR = 0.72, 95% CI = 0.54-0.97, P = 0.03) and the ratio cer16/cer24:0 (HR =1.32, 95% CI = 1.04-1.67, P = 0.02) were statistically significant in the fully adjusted models, but not in the crude model.

In the model with kidney failure as outcome, cer16 and ratios cer20/cer24:0, cer22/cer24:0 and cer24:1/cer24:0 were significantly associated in crude models. In the level 1 adjusted model all the ratios to cer24:0 were significantly associated with kidney failure (HR =2.1, 95% CI =1.37-3.22, P =6.56*10^−4^), (HR =1.69, 95% CI =1.11-2.58, P =0.01), (HR =1.73, 95% CI =1.14-2.62, P =0.01), (HR =1.9, 95% CI =1.23-2.94, P =3.83*10^−3^),(HR =2.06, 95% CI =1.38-3.07, P =4.14*10^−4^) for cer16/cer24:0, cer18/cer24:0, cer20/cer24:0, cer22/cer24:0 and cer24:1/cer24:0 respectively, however none of these results persisted after inclusion of eGFR and UAER in the model. A full list of model estimates for all the models and adjustment levels can be found in Supplementary Table 1 with a visual summary in Figure 1.

Cer20, cer22, cer24:0 and cer24:1 were all significantly inversely associated with all-cause mortality and this finding persisted through all three levels of adjustments. The strongest metabolite associations to all-cause mortality in the adjusted models were cer22 (hazard ratio (HR) = 0.38, 95% CI = 0.24-0.60, P = 3.3*10^−5^) and cer24:0 (HR = 0.47, 95% CI = 0.31-0.73, P = 6.6*10^−4^). The ratios cer16/cer24:0 and cer18/cer24:0 were also associated with all-cause mortality throughout all levels of adjustments (HR = 1.69, 95% CI = 1.27-2.23, P = 2.8*10^−4^) and (HR = 1.48, 95% CI = 1.12-1.94, P = 5*10^−3^ respectively in the fully adjusted model).

The ratio of cer18/cer24:0 was the only measure that was significantly associated with CVE and all-cause mortality at all levels of adjustments. Kaplan-Meier curves (Figure 2) show that participants above the median of ratio of cer18/cer24:0 were at higher risk (81% survival rate, 95% CI = 77-85, six years after measuring) than participants below the median (89% survival rate, 95% CI = 85-92, six years after measuring, P = 0.013) for CVE. The ceramide ratio cer18/cer24:0 was a better at differentiating CVE from non-CVE than LDL, based on which participants above the median of LDL were at a similar risk (84% survival rate, 95% CI = 0.80-0.88 six years after measuring) to the participants with LDL lower than the median (87% survival rate, 95% CI = 0.83-0.91 six years after measuring, P = 0.2).

**Figure 2.**
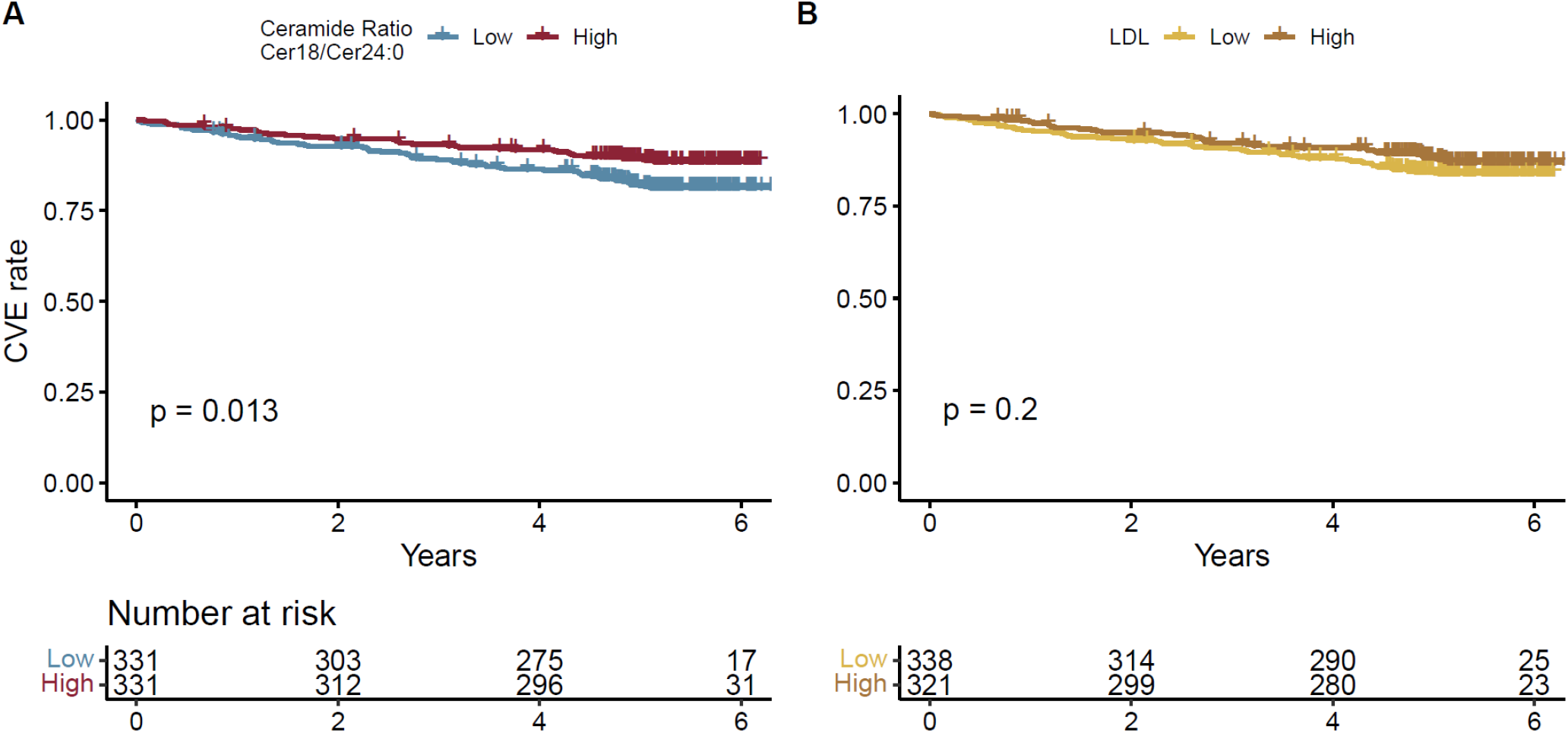
Kaplan Meier plot of cer18/cer24:0 ratio and LDL against CVE. High is metabolite level greater than or equal to the median, low is individuals with a metabolite level below the median.

Examining the variables used for adjustment we found that CVD history was the variable with the strongest association with CVE (HR = 3.72, 95% CI =2.34-5.92, P =2.84*10^−8^), Supplementary Figure 1 and Supplementary Table 3. The ceramides showed strongest correlation with each other and with the other lipid measures like LDL and triglycerides correlation coefficients between 0.3-45 and 0.27-0.5 respectively, Figure 3, extended heatmap in Supplementary Figure 2. In addition, cer24:0 had a small positive correlation with eGFR (0.08), compared to the other ceramides that had negative or no correlation with eGFR.

**Figure 3.**
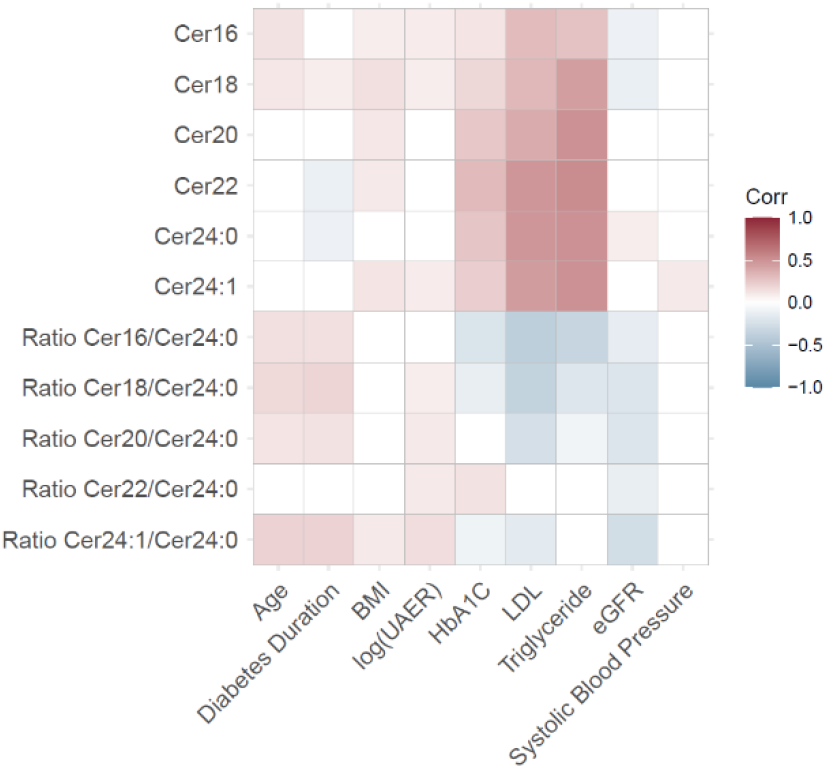
Heatmap of ceramides correlation to possible CVE confounders, presented as correlations coefficients from Pearson’s correlation.

Finally, we investigated how the HRs for CVE were affected by albuminuria and found that from normoalbuminuria to moderate increase in albuminuria cer16 and cer18 were significantly associated with CVE, but in the subpopulation with severely increased albuminuria all significant associations were lost (Figure 4). The levels of cer18, cer24:0 and the ratio of cer18/cer24:0 were plotted for each albuminuria group and for CVE (Supplementary Figure 3) which shows that the difference in ceramide level between individuals with and without a CVE is gradually smaller with worsening albuminuria.

**Figure 4.**
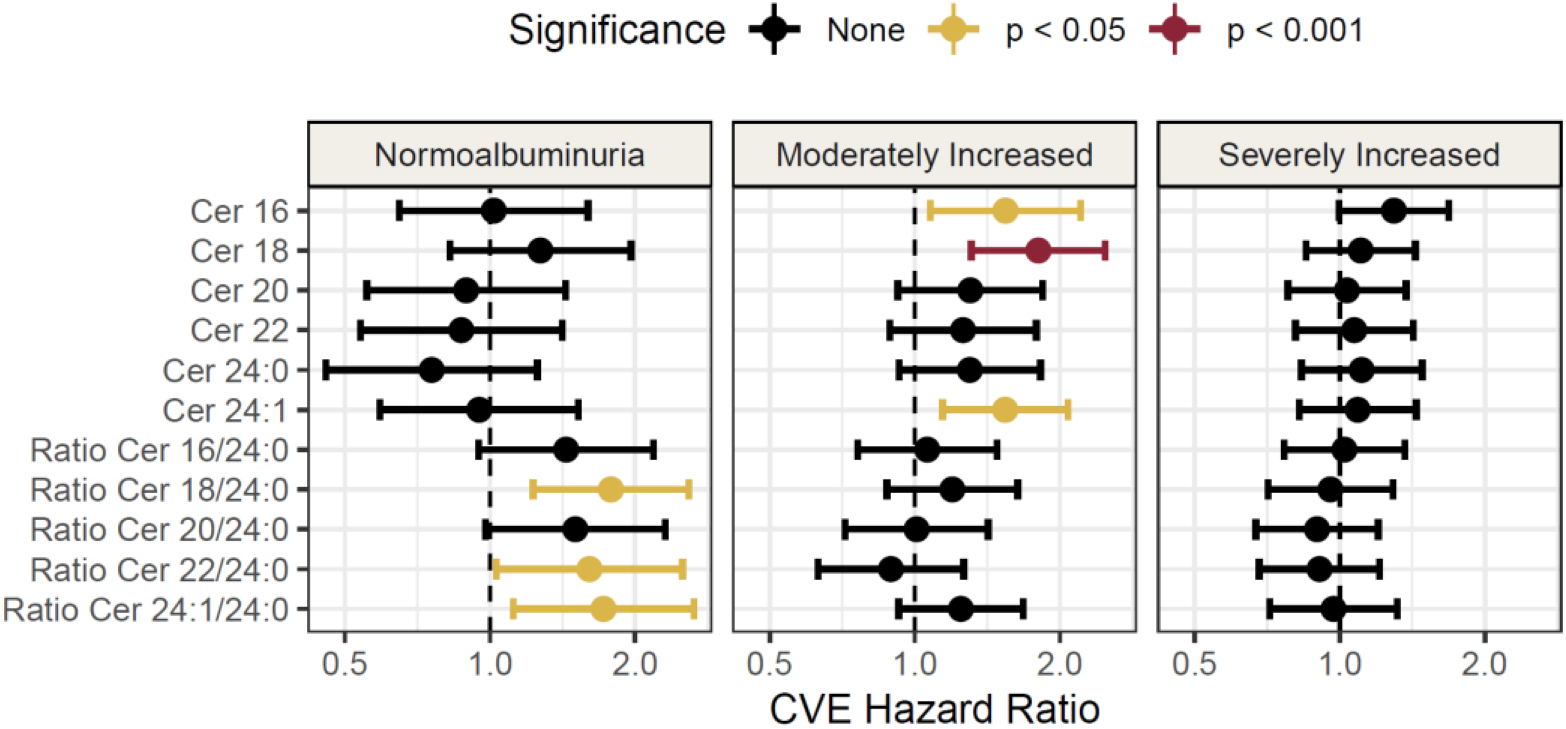
Forest plot of hazard ratios for ceramide and ratios for cardiovascular events separated by albuminuria status. Normoalbuminuria is defined as <30 mg/g, moderately increased is between 30-299 mg/g and severely increased is >=300 mg/g. These models are unadjusted crude models. Hazard ratios (HRs) are reported per doubling of the log10 ceramide.

## Discussion

We investigated the potential of a small set of specific circulating ceramides as biomarkers for cardiovascular risk, kidney failure and all-cause mortality in individuals with T1D. In brief, the key findings are: (1) Several of the investigated ceramides associated with CVE risk and all-cause mortality in individuals with T1D. (2) The cer18/cer24:0 ratio was the most consistent measure and showed constant, significant, association with CVE and all-cause mortality across all confounder adjustment levels. The cer18/cer24:0 ratio furthermore outperformed LDL cholesterol in separating individuals who progressed to a CVE from the group who did not. (3) Ratios of the ceramide cer24:0 associated, albeit weaker, to kidney failure, until the adjustment for UAER and eGFR. The loss of association after adjustment to UAER and eGFR may be explained by collinearity between ceramide ratios and eGFR (Figure 3) (4). We observed that the ceramides ability to differentiate between individuals with and without future CVE depended on albuminuria status; whether this is a direct effect or an indicator of compounding illness is not clear, though it should be noted that these ceramides have previously been associated with albuminuria and eGFR (30–32).

There is compelling evidence that ceramides can predict CVD risk. Tarasov *et al*. 2014 were among the first to report increased levels of cer16, cer18, cer20 and cer24:1 as risk markers of coronary artery disease together with a reduced risk associated with cer24:0 (9). Several subsequent studies found associations for ceramide levels and cardiovascular risk (33–35), however, few have studied these biomarkers in relation to diabetes. Alshehry *et al*. investigated lipids associated with CVE and cardiovascular death in a T2D cohort and found, among other lipids, that cer24:1 was positively associated with CVE and cardiovascular death (36). We did not observe an association between cer24:1 and CVE, however we did observe a protective association to all-cause mortality. We have not been able to find any studies investigating the cardiovascular risk of these ceramides in people with T1D. Lipid measures are well established measures of CVD risk (37,38). We found that the ratio of cer18/cer24:0 could be a comparable measure to LDL. Havulinna *et al*. showed that adding cer18 measures to LDL levels improved their ability to predict incident rate of cardiovascular events, while adding LDL levels to cer18 level did not improve prediction (11).

The association between ceramides and kidney disease in the literature is a less clear. One study found that the amount of cer20 and cer24:1 lowered with worsening albuminuria in individuals with T1D (30), which could explain why their ability to distinguish CVE from non-CVE could be hampered in individuals with severe kidney disease. Contradicting this, have studies found increase in ceramide levels in people with chronic kidney disease (32) and diabetic kidney disease (31) compared to controls without kidney disease, the increase of cer16 found in these two studies, matches with the association between cer16 and risk of kidney failure found in this study (Figure 1). Considering these reports and our study, it is possible that ceramide levels increase with kidney/endothelial damage up until a tipping point, where the levels then drop; a similar metabolic tipping point have been suggested in individuals with severe liver fibrosis (39). The influence of albuminuria status on ceramides ability to associate with CVE should be investigated further and be considered when using these ceramides as risk predictors.

This study has some limitations. We were not able to control for diet, exercise and change in medication in this post-hoc analysis, which could have influenced ceramide levels (12,18). It should be noted that the participants had longstanding T1D and therefore were likely to adhere low-carb diet without major changes. Despite the possible viability introduced by uncontrolled factors, we found robust associations between ceramides, CVE and all-cause mortality. Validating our finding in an independent replication cohort, would help cement our results further, however we were not able to identify a similar cohort. For the strengths, the levels of the ceramides were measured in a semiquantitative manner allowing for comparison with other future studies. Additionally, investigating few specific molecular targets measured in concentrations reduce the risk of incidental discoveries. It was also a strength that this analysis was caried out in a large well characterized prospective cohort.

In this study we investigated a small set of specific ceramides in relation to CVE, kidney failure and all-cause mortality. We found that these ceramides have the potential as susceptibility biomarkers of CVD and all-cause mortality in individuals with longstanding T1D. Cer18/cer24:0 appear to be the strongest, independent risk marker for CVD and all-cause mortality, and future work should focus on validating it in clinical practice.

## Data Availability

The dataset analyzed here is not publicly available, for the privacy of the participants, in compliance with EU and Danish data protection law. The data can be accessed upon request; relevant legal permission from the data protection agency is required. Data access request should be directed to PR.
The code used for data analysis is available on github: https://github.com/Asger-W/Profil-Ceramides.

## List of abbreviations

BMI: Body mass index
Cer: Ceramide
CI: Confidence interval
CVD: Cardiovascular disease
CVE: Cardiovascular events
eGFR: Estimated glomerular filtration rate
HbA_1c_: Hemoglobin A1C
HR: Hazard Ratio
LDL: Low-density lipids
T1D: Type 1 Diabetes
UAER: Urinary albumin excretion rate
VLDL: Very low-density lipids.

## Declarations

### Ethics approval and consent to participate

The original study was approved by the Danish National Committee on Biomedical Research Ethics (2009-056). The follow-up protocol was approved The Ethics Committee E, Region Hovedstaden, Denmark. Both the original study and the follow-up protocol adheres to principles of the Declaration of Helsinki. Written and informed consent was given by the participants before inclusion in the original study, which included consent for inclusion in a follow up study.

### Availability of data and materials

The dataset analyzed here is not publicly available, for the privacy of the participants, in compliance with EU and Danish data protection law. The data can be accessed upon request; relevant legal permission from the data protection agency is required. Data access request should be directed to PR.

The code used for data analysis is available on github: https://github.com/Asger-W/Profil-Ceramides.

### Authors’ contributions

AW performed data analysis and drafted the manuscript. AW, VRC, TS, ST, NT, SAW, TV, HV PR and CLQ contributed to the conceptualization and interpretation of this study, ST and PR conducted the original study, VRC, TS, NT and SAW carried out the follow-up study and provided material and clinical data for this study.

## References

1. Patterson CC, Karuranga S, Salpea P, Saeedi P, Dahlquist G, Soltesz G, et al. Worldwide estimates of incidence, prevalence and mortality of type 1 diabetes in children and adolescents: Results from the International Diabetes Federation Diabetes Atlas, 9th edition. Diabetes Res Clin Pract [Internet]. 2019;157:107842. Available from: https://doi.org/10.1016/j.diabres.2019.107842

2. Jørgensen ME, Almdal TP, Carstensen B. Time trends in mortality rates in type 1 diabetes from 2002 to 2011. Diabetologia. 2013;56(11):2401–4.

3. Rawshani A, Sattar N, Franzén S, Rawshani A, Hattersley AT, Svensson AM, et al. Excess mortality and cardiovascular disease in young adults with type 1 diabetes in relation to age at onset: a nationwide, register-based cohort study. Lancet. 2018;392(10146):477–86.

4. Bjerg L, Hulman A, Carstensen B, Charles M, Witte DR, Jørgensen ME. Effect of duration and burden of microvascular complications on mortality rate in type 1 diabetes: an observational clinical cohort study. Diabetologia. 2019;41(11):2297–305.

5. de Boer IH, Gao X, Cleary PA, Bebu I, Lachin JM, Molitch ME, et al. Albuminuria changes and cardiovascular and renal outcomes in type 1 diabetes: The DCCT/EDIC study. Clin J Am Soc Nephrol. 2016;11(11):1969–77.

6. Lees JS, Welsh CE, Celis-Morales CA, Mackay D, Lewsey J, Gray SR, et al. Glomerular filtration rate by differing measures, albuminuria and prediction of cardiovascular disease, mortality and end-stage kidney disease. Nat Med [Internet]. 2019;25(11):1753–60. Available from: http://dx.doi.org/10.1038/s41591-019-0627-8

7. Dennis JM. Precision medicine in type 2 diabetes: Using individualized prediction models to optimize selection of treatment. Diabetes. 2020;69(10):2075–85.

8. Buse JB, Wexler DJ, Tsapas A, Rossing P, Mingrone G, Mathieu C, et al. 2019 Update to: Management of Hyperglycemia in Type 2 Diabetes, 2018. A Consensus Report by the American Diabetes Association (ADA) and the European Association for the Study of Diabetes (EASD). Diabetes Care. 2020;43(February):487–93.

9. Tarasov K, Ekroos K, Suoniemi M, Kauhanen D, Sylvänne T, Hurme R, et al. Molecular lipids identify cardiovascular risk and are efficiently lowered by simvastatin and PCSK9 deficiency. J Clin Endocrinol Metab. 2014;99(1):45–52.

10. Laaksonen R, Ekroos K, Sysi-Aho M, Hilvo M, Vihervaara T, Kauhanen D, et al. Plasma ceramides predict cardiovascular death in patients with stable coronary artery disease and acute coronary syndromes beyond LDL-cholesterol. Eur Heart J. 2016;37(25):1967–76.

11. Havulinna AS, Sysi-Aho M, Hilvo M, Kauhanen D, Hurme R, Ekroos K, et al. Circulating Ceramides Predict Cardiovascular Outcomes in the Population-Based FINRISK 2002 Cohort. Arterioscler Thromb Vasc Biol. 2016;36(12):2424–30.

12. Wang DD, Toledo E, Hruby A, Rosner BA, Willett WC, Sun Q, et al. Plasma ceramides, mediterranean diet, and incident cardiovascular disease in the PREDIMED trial (prevención con dieta mediterránea). Circulation. 2017;135(21):2028–40.

13. Peterson LR, Xanthakis V, Duncan MS, Gross S, Friedrich N, Völzke H, et al. Ceramide remodeling and risk of cardiovascular events and mortality. J Am Heart Assoc. 2018;7(10).

14. Lemaitre RN, Jensen PN, Hoofnagle A, Mcknight B, Fretts AM, King IB, et al. Plasma Ceramides and Sphingomyelins in Relation to Heart Failure Risk: The Cardiovascular Health Study. Circ Hear Fail. 2019;12(7):1–8.

15. Choi RH, Tatum SM, Symons JD, Summers SA, Holland WL. Ceramides and other sphingolipids as drivers of cardiovascular disease. Nat Rev Cardiol [Internet]. 2021;18(10):701–11. Available from: http://dx.doi.org/10.1038/s41569-021-00536-1

16. Haus JM, Kashyap SR, Kasumov T, Zhang R, Kelly KR, Defronzo RA, et al. Plasma ceramides are elevated in obese subjects with type 2 diabetes and correlate with the severity of insulin resistance. Diabetes. 2009;58(2):337–43.

17. Huang H, Kasumov T, Gatmaitan P, Heneghan HM, Kashyap SR, Schauer PR, et al. Gastric bypass surgery reduces plasma ceramide subspecies and improves insulin sensitivity in severely obese patients. Obesity [Internet]. 2011;19(11):2235–40. Available from: http://dx.doi.org/10.1038/oby.2011.107/nature06264

18. Bergman BC, Brozinick JT, Strauss A, Bacon S, Kerege A, Bui HH, et al. Serum sphingolipids: Relationships to insulin sensitivity and changes with exercise in humans. Am J Physiol - Endocrinol Metab. 2015;309(4):E398–408.

19. Lemaitre RN, Yu C, Hoofnagle A, Hari N, Jensen PN, Fretts AM, et al. Circulating sphingolipids, insulin, HOMA-IR, and HOMA-B: The Strong heart family study. Diabetes. 2018;67(8):1663–72.

20. Hilvo M, Salonurmi T, Havulinna AS, Kauhanen D, Pedersen ER, Tell GS, et al. Ceramide stearic to palmitic acid ratio predicts incident diabetes. Diabetologia. 2018;61(6):1424–34.

21. Chew WS, Torta F, Ji S, Choi H, Begum H, Sim X, et al. Large-scale lipidomics identifies associations between plasma sphingolipids and T2DM incidence. JCI Insight. 2019;4(13):1–14.

22. Fretts AM, Jensen PN, Hoofnagle A, McKnight B, Howard B V., Umans J, et al. Plasma ceramide species are associated with diabetes risk in participants of the strong heart study. J Nutr. 2020;150(5):1214–22.

23. Theilade S, Lajer M, Hansen TW, Joergensen C, Persson F, Andrésdottir G, et al. 24-Hour Central Aortic Systolic Pressure and 24-Hour Central Pulse Pressure Are Related To Diabetic Complications in Type 1 Diabetes - a Cross-Sectional Study. Cardiovasc Diabetol. 2013;12(1).

24. Tofte N, Suvitaival T, Ahonen L, Winther SA, Theilade S, Frimodt-møller M, et al. Lipidomic analysis reveals sphingomyelin and phosphatidylcholine species associated with renal impairment and all-cause mortality in type 1 diabetes. Sci Rep. 2019;1–10.

25. Curovic VR, Suvitaival T, Mattila I, Ahonen L, Tro K, Theilade S, et al. Circulating Metabolites and Lipids Are Associated to Diabetic Retinopathy in Individuals With Type 1 Diabetes. Diabetes. 2020;69(July):2217–26.

26. Pluskal T, Castillo S, Villar-briones A, Ore M. MZmine 2: Modular framework for processing, visualizing, and analyzing mass spectrometry-based molecular profile data. BMC Bioinformatics. 2010;11(395):1–11.

27. Kim M, Nevado-Holgado A, Whiley L, Snowden SG, Soininen H, Kloszewska I, et al. Association between Plasma Ceramides and Phosphatidylcholines and Hippocampal Brain Volume in Late Onset Alzheimer’s Disease. J Alzheimers Dis. 2017;60(3):809–17.

28. R Core Team (2018). R: A language and environment for statistical computing. [Internet]. Vienna, Austria.: R Foundation for Statistical Computing; 2018. Available from: https://www.r-project.org/.

29. Divino F, Divino LFF, Suvitaival T, Curovic VR, Tofte N, Trošt K, et al. Circulating metabolites and molecular lipid species are associated with future cardiovascular morbidity and mortality in type 1 diabetes. Cardiovasc Diabetol [Internet]. 2022;1–10. Available from: https://doi.org/10.1186/s12933-022-01568-8

30. Klein RL, Hammad SM, Baker NL, Hunt KJ, Al Gadban MM, Cleary PA, et al. Decreased plasma levels of select very long chain ceramide species Are associated with the development of nephropathy in type 1 diabetes. Metabolism [Internet]. 2014;63(10):1287–95. Available from: http://dx.doi.org/10.1016/j.metabol.2014.07.001

31. Liu JJ, Ghosh S, Kovalik JP, Ching J, Choi HW, Tavintharan S, et al. Profiling of Plasma Metabolites Suggests Altered Mitochondrial Fuel Usage and Remodeling of Sphingolipid Metabolism in Individuals With Type 2 Diabetes and Kidney Disease. Kidney Int Reports [Internet]. 2017;2(3):470–80. Available from: http://dx.doi.org/10.1016/j.ekir.2016.12.003

32. Mantovani A, Lunardi G, Bonapace S, Dugo C, Altomari A, Molon G, et al. Association between increased plasma ceramides and chronic kidney disease in patients with and without ischemic heart disease. Diabetes Metab [Internet]. 2021;47(1):101152. Available from: https://doi.org/10.1016/j.diabet.2020.03.003

33. Cheng JM, Suoniemi M, Kardys I, Vihervaara T, de Boer SPM, Akkerhuis KM, et al. Plasma concentrations of molecular lipid species in relation to coronary plaque characteristics and cardiovascular outcome: Results of the ATHEROREMO-IVUS study. Atherosclerosis [Internet]. 2015;243(2):560–6. Available from: http://dx.doi.org/10.1016/j.atherosclerosis.2015.10.022

34. Anroedh S, Hilvo M, Martijn Akkerhuis K, Kauhanen D, Koistinen K, Oemrawsingh R, et al. Plasma concentrations of molecular lipid species predict long-term clinical outcome in coronary artery disease patients. J Lipid Res [Internet]. 2018;59(9):1729–37. Available from: http://dx.doi.org/10.1194/jlr.P081281

35. Meeusen JW, Donato LJ, Bryant SC, Baudhuin LM, Berger PB, Jaffe AS. Plasma ceramides a novel predictor of major adverse cardiovascular events after coronary angiography. Arterioscler Thromb Vasc Biol. 2018;38(8):1933–9.

36. Alshehry ZH, Mundra PA, Barlow CK, Mellett NA, Wong G, McConville MJ, et al. Plasma Lipidomic Profiles Improve on Traditional Risk Factors for the Prediction of Cardiovascular Events in Type 2 Diabetes Mellitus. Circulation. 2016;134(21):1637–50.

37. Iso H, Jacobs DR, Wentworth D, Neaton JD, Cohen JD. Serum Cholesterol Levels and Six-Year Mortality From Stroke in 350,977 Men Screened for the Multiple Risk Factor Intervention Trial. N Engl J Med. 1989;320(14):904–10.

38. Ference BA, Ginsberg HN, Graham I, Ray KK, Chris J, Bruckert E, et al. Low-density lipoproteins cause atherosclerotic cardiovascular disease. 1. Evidence from genetic, epidemiologic, and clinical studies. A consensus statement from the European Atherosclerosis Society Consensus Panel. Eur Heart J. 2017;38:2459–72.

39. McGlinchey AJ, Govaere O, Geng D, Ratziu V, Allison M, Bousier J, et al. Metabolic signatures across the full spectrum of non-alcoholic fatty liver disease. JHEP Reports. 2022;4(5).

