## Supplementary data for "Ceramides as risk markers for future cardiovascular events and all-cause mortality in long-standing type 1 diabetes"

Supplementary figure 1: Forest plot of hazard ratios for clinical variables for outcomes of cardiovascular events, kidney failure and mortality

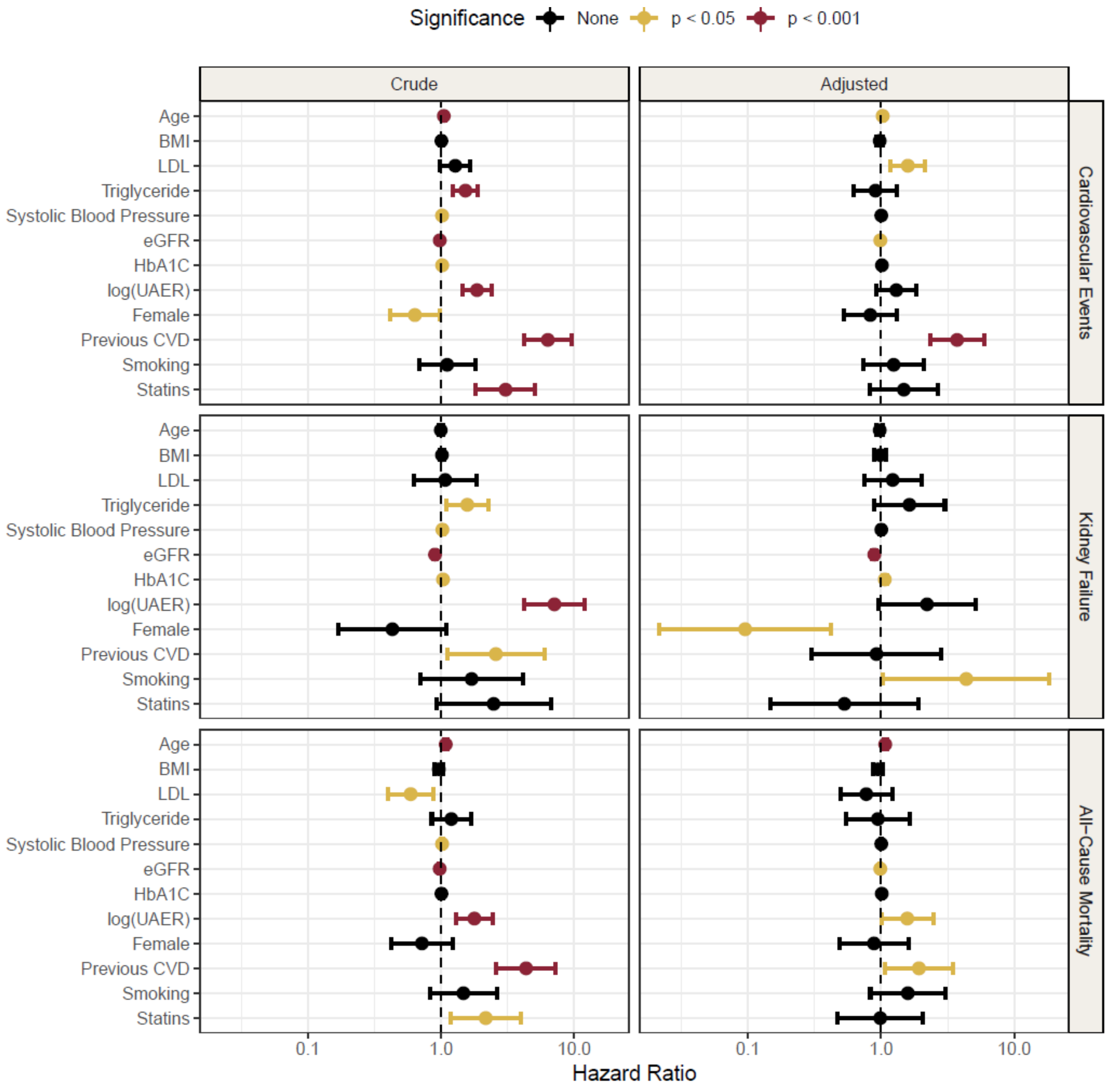


Survival analysis of clinical variables against cardiovascular events, kidney failure and mortality. The crude models are unadjusted. The adjusted model is adjusted all the other clinical variables than the one interrogated. Hazard ratios are reported per doubling of the variable in question.

Supplementary figure 2: Correlation matrix with correlation coefficients

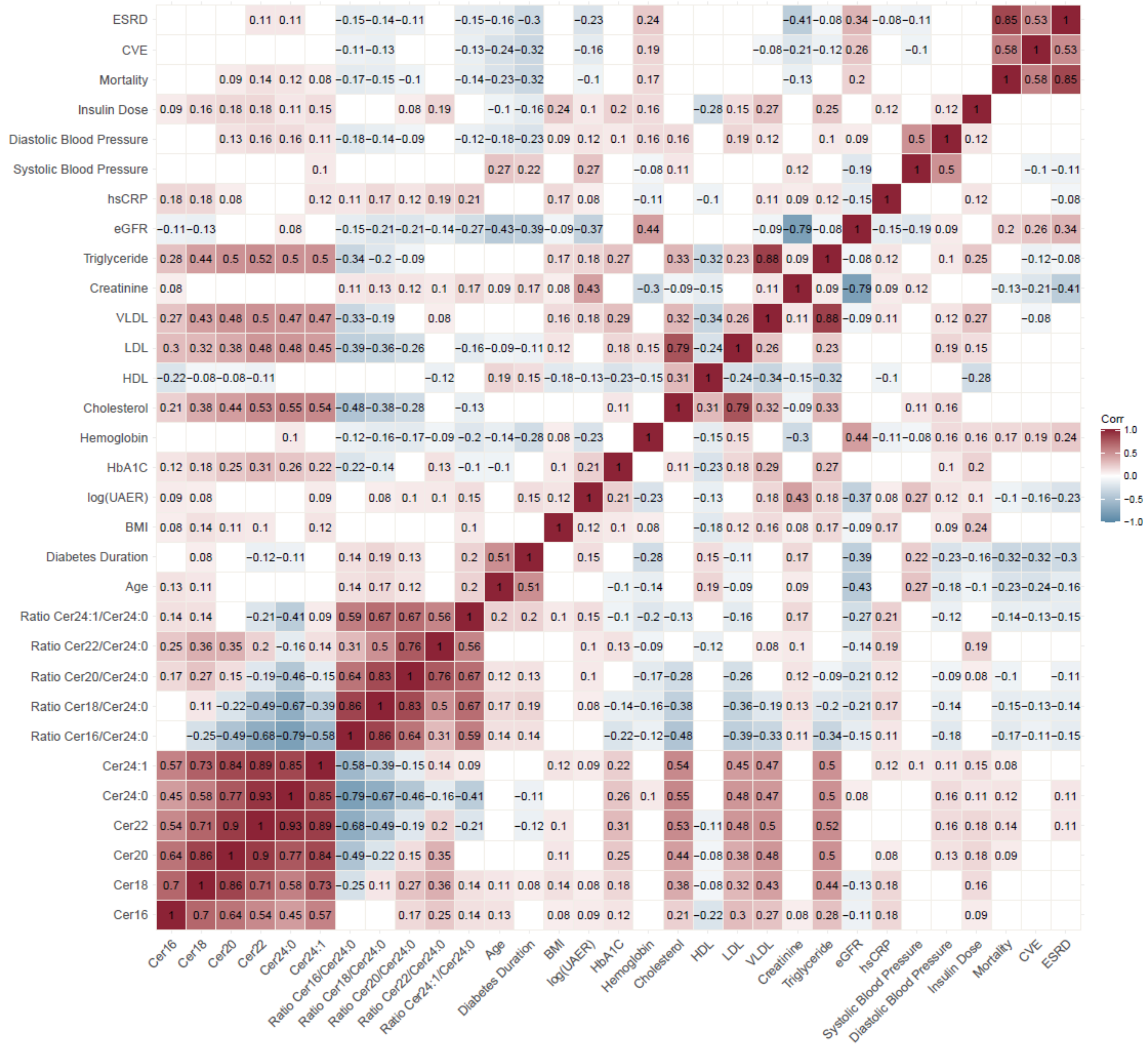


The values indicate the correlations coefficients from Pearson correlations, only significant correlations shown.

Supplementary figure 3: Boxplot of ratio cer18/cer24:0 and its constituents distributed by CVE- and albuminuria status.

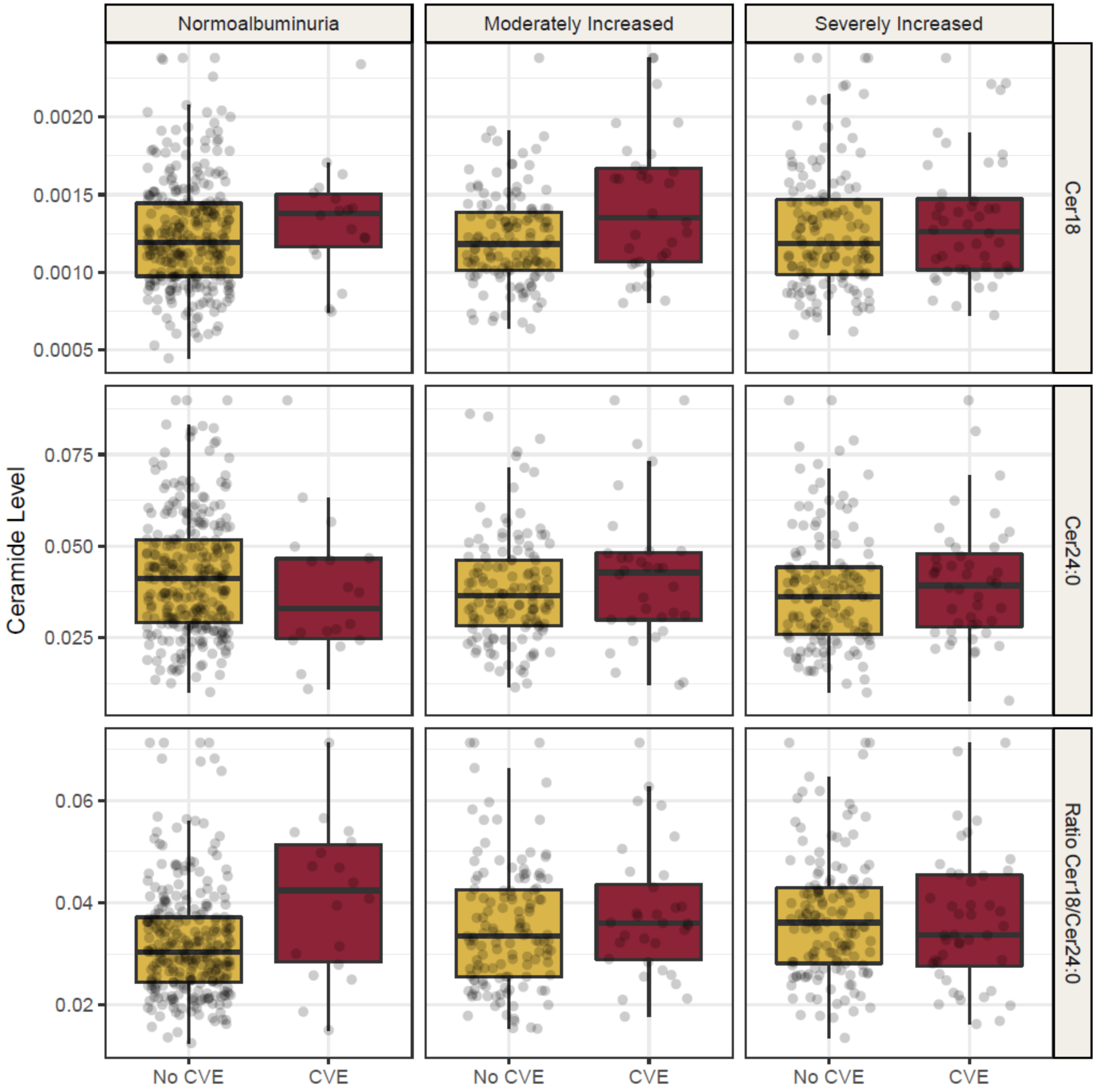

Supplementary table 1: Ceramide quartiles

| | Min. | Q1 | Median | Q3 | Max |
|---|---|---|---|---|---|
| Cer16 ng/ml | 0.86 | 1.91 | 2.18 | 2.48 | 3.58 |
| Cer18 ng/ml | 0.45 | 0.99 | 1.20 | 1.45 | 2.38 |
| Cer20 ng/ml | 0.42 | 1.31 | 1.69 | 2.14 | 3.75 |
| Cer22 ng/ml | 2.17 | 7.10 | 9.6 | 12.57 | 23.10 |
| Cer24:0 ng/ml | 7.78 | 28.00 | 37.75 | 48.93 | 89.78 |
| Cer24:1 ng/ml | 5.38 | 14.08 | 18.50 | 23.20 | 41.82 |
| Ratio Cer16/Cer24:0 | 0.01 | 0.05 | 0.06 | 0.07 | 0.14 |
| Ratio Cer18/Cer24:0 | 0.01 | 0.03 | 0.03 | 0.04 | 0.07 |
| Ratio Cer20/Cer24:0 | 0.02 | 0.04 | 0.05 | 0.05 | 0.08 |
| Ratio Cer22/Cer24:0 | 0.15 | 0.23 | 0.26 | 0.28 | 0.37 |
| Ratio Cer24:1/Cer24:0 | 0.27 | 0.43 | 0.49 | 0.57 | 0.83 |

Supplementary table 2: Hazard ratios of ceramides and ratios for outcomes of cardiovascular events, kidney disease and all-cause mortality

| Name | Hazard Ratio | CI lower | CI upper | p-value | Model | Outcome |
|---|---|---|---|---|---|---|
| Cer16 | 1.287044 | 1.061995 | 1.559784 | 0.010072 | Crude | CVE |
| Cer18 | 1.324777 | 1.102941 | 1.591232 | 0.002631 | Crude | CVE |
| Cer20 | 1.061712 | 0.871102 | 1.29403 | 0.553092 | Crude | CVE |
| Cer22 | 1.030734 | 0.843064 | 1.260181 | 0.767838 | Crude | CVE |
| Cer24:0 | 1.012705 | 0.82557 | 1.242258 | 0.903599 | Crude | CVE |
| Cer24:1 | 1.196914 | 0.991062 | 1.445524 | 0.06194 | Crude | CVE |
| Ratio cer16/cer24:0 | 1.193681 | 0.987117 | 1.443471 | 0.067819 | Crude | CVE |
| Ratio cer18/cer24:0 | 1.29242 | 1.076807 | 1.551205 | 0.005876 | Crude | CVE |
| Ratio cer20/cer24:0 | 1.138701 | 0.934594 | 1.387384 | 0.197472 | Crude | CVE |
| Ratio cer22/cer24:0 | 1.076939 | 0.879711 | 1.318385 | 0.472643 | Crude | CVE |
| Ratio cer24:1/cer24:0 | 1.340615 | 1.114388 | 1.612768 | 0.00188 | Crude | CVE |
| Cer16 | 1.038481 | 0.832095 | 1.296057 | 0.738371 | Level 2 adjusted | CVE |
| Cer18 | 1.034406 | 0.818786 | 1.306809 | 0.776697 | Level 2 adjusted | CVE |
| Cer20 | 0.76927 | 0.584382 | 1.012654 | 0.061441 | Level 2 adjusted | CVE |
| Cer22 | 0.721673 | 0.536581 | 0.970611 | 0.030987 | Level 2 adjusted | CVE |
| Cer24:0 | 0.760712 | 0.571693 | 1.012227 | 0.060576 | Level 2 adjusted | CVE |
| Cer24:1 | 0.832043 | 0.630213 | 1.09851 | 0.194581 | Level 2 adjusted | CVE |
| Ratio cer16/cer24:0 | 1.320287 | 1.040819 | 1.674794 | 0.022041 | Level 2 adjusted | CVE |
| Ratio cer18/cer24:0 | 1.334997 | 1.06017 | 1.681068 | 0.014018 | Level 2 adjusted | CVE |
| Ratio cer20/cer24:0 | 1.084142 | 0.861519 | 1.364293 | 0.490881 | Level 2 adjusted | CVE |
| Ratio cer22/cer24:0 | 0.962719 | 0.767451 | 1.20767 | 0.742533 | Level 2 adjusted | CVE |
| Ratio cer24:1/cer24:0 | 1.098969 | 0.885945 | 1.363215 | 0.390658 | Level 2 adjusted | CVE |
| Cer16 | 1.056322 | 0.849842 | 1.312968 | 0.621474 | Level 1 adjusted | CVE |
| Cer18 | 1.039419 | 0.825096 | 1.309414 | 0.742797 | Level 1 adjusted | CVE |
| Cer20 | 0.776127 | 0.595835 | 1.010972 | 0.060237 | Level 1 adjusted | CVE |
| Cer22 | 0.734233 | 0.552747 | 0.975308 | 0.032961 | Level 1 adjusted | CVE |
| Cer24:0 | 0.754939 | 0.573472 | 0.993827 | 0.045059 | Level 1 adjusted | CVE |

| | | | | | | |
|---|---|---|---|---|---|---|
| Cer24:1 | 0.83481 | 0.638957 | 1.090696 | 0.185652 | Level 1 adjusted | CVE |
| Ratio cer16/cer24:0 | 1.368911 | 1.085914 | 1.725659 | 0.007872 | Level 1 adjusted | CVE |
| Ratio cer18/cer24:0 | 1.373097 | 1.096159 | 1.720001 | 0.005801 | Level 1 adjusted | CVE |
| Ratio cer20/cer24:0 | 1.118899 | 0.893883 | 1.400557 | 0.32674 | Level 1 adjusted | CVE |
| Ratio cer22/cer24:0 | 1.018689 | 0.818111 | 1.268443 | 0.868546 | Level 1 adjusted | CVE |
| Ratio cer24:1/cer24:0 | 1.137629 | 0.922604 | 1.402769 | 0.227686 | Level 1 adjusted | CVE |
| Cer16 | 1.518718 | 1.041621 | 2.214342 | 0.029862 | Crude | Kidney failure |
| Cer18 | 1.200416 | 0.822554 | 1.75186 | 0.343574 | Crude | Kidney failure |
| Cer20 | 1.162892 | 0.793799 | 1.703602 | 0.438564 | Crude | Kidney failure |
| Cer22 | 1.134795 | 0.772637 | 1.666707 | 0.519089 | Crude | Kidney failure |
| Cer24:0 | 0.914094 | 0.598628 | 1.395804 | 0.677482 | Crude | Kidney failure |
| Cer24:1 | 1.308074 | 0.911362 | 1.877473 | 0.145237 | Crude | Kidney failure |
| Ratio cer16/cer24:0 | 1.421261 | 0.997994 | 2.024041 | 0.051315 | Crude | Kidney failure |
| Ratio cer18/cer24:0 | 1.336519 | 0.92903 | 1.922739 | 0.117996 | Crude | Kidney failure |
| Ratio cer20/cer24:0 | 1.514159 | 1.043582 | 2.196931 | 0.028918 | Crude | Kidney failure |
| Ratio cer22/cer24:0 | 1.873259 | 1.273066 | 2.756416 | 0.001447 | Crude | Kidney failure |
| Ratio cer24:1/cer24:0 | 1.788425 | 1.262751 | 2.532935 | 0.001061 | Crude | Kidney failure |
| Cer16 | 0.785812 | 0.426414 | 1.448125 | 0.439634 | Level 2 adjusted | Kidney failure |
| Cer18 | 0.763994 | 0.414952 | 1.406639 | 0.387382 | Level 2 adjusted | Kidney failure |
| Cer20 | 0.783169 | 0.411842 | 1.489294 | 0.456074 | Level 2 adjusted | Kidney failure |
| Cer22 | 0.88226 | 0.442041 | 1.760884 | 0.722388 | Level 2 adjusted | Kidney failure |
| Cer24:0 | 0.746554 | 0.392017 | 1.421731 | 0.373826 | Level 2 adjusted | Kidney failure |
| Cer24:1 | 1.313844 | 0.605066 | 2.852888 | 0.490213 | Level 2 adjusted | Kidney failure |
| Ratio cer16/cer24:0 | 1.231547 | 0.639975 | 2.36995 | 0.532893 | Level 2 adjusted | Kidney failure |
| Ratio cer18/cer24:0 | 1.02778 | 0.590414 | 1.789137 | 0.922819 | Level 2 adjusted | Kidney failure |
| Ratio cer20/cer24:0 | 1.176961 | 0.64689 | 2.141381 | 0.593641 | Level 2 adjusted | Kidney failure |

| | | | | | | |
|---|---|---|---|---|---|---|
| Ratio cer22/cer24:0 | 1.517269 | 0.840991 | 2.737371 | 0.166124 | Level 2 adjusted | Kidney failure |
| Ratio cer24:1/cer24:0 | 1.516774 | 0.902093 | 2.550296 | 0.116109 | Level 2 adjusted | Kidney failure |
| Cer16 | 1.422956 | 0.910476 | 2.223895 | 0.121551 | Level 1 adjusted | Kidney failure |
| Cer18 | 1.00652 | 0.634482 | 1.596709 | 0.977977 | Level 1 adjusted | Kidney failure |
| Cer20 | 0.908924 | 0.560147 | 1.474868 | 0.699012 | Level 1 adjusted | Kidney failure |
| Cer22 | 0.806301 | 0.475126 | 1.368314 | 0.424944 | Level 1 adjusted | Kidney failure |
| Cer24:0 | 0.590765 | 0.342427 | 1.019208 | 0.058546 | Level 1 adjusted | Kidney failure |
| Cer24:1 | 1.038422 | 0.643272 | 1.676305 | 0.877369 | Level 1 adjusted | Kidney failure |
| Ratio cer16/cer24:0 | 2.100597 | 1.370627 | 3.219335 | 0.000656 | Level 1 adjusted | Kidney failure |
| Ratio cer18/cer24:0 | 1.690377 | 1.107379 | 2.580304 | 0.01499 | Level 1 adjusted | Kidney failure |
| Ratio cer20/cer24:0 | 1.728001 | 1.14017 | 2.618898 | 0.009928 | Level 1 adjusted | Kidney failure |
| Ratio cer22/cer24:0 | 1.900115 | 1.229733 | 2.935952 | 0.003834 | Level 1 adjusted | Kidney failure |
| Ratio cer24:1/cer24:0 | 2.059266 | 1.379055 | 3.074987 | 0.000414 | Level 1 adjusted | Kidney failure |
| Cer16 | 1.070482 | 0.830049 | 1.380558 | 0.599742 | Crude | Mortality |
| Cer18 | 0.925248 | 0.710124 | 1.205543 | 0.564992 | Crude | Mortality |
| Cer20 | 0.728085 | 0.544482 | 0.9736 | 0.032321 | Crude | Mortality |
| Cer22 | 0.54495 | 0.391647 | 0.75826 | 0.000316 | Crude | Mortality |
| Cer24:0 | 0.583685 | 0.425187 | 0.801267 | 0.000867 | Crude | Mortality |
| Cer24:1 | 0.734411 | 0.547006 | 0.98602 | 0.040012 | Crude | Mortality |
| Ratio cer16/cer24:0 | 1.765313 | 1.43304 | 2.174627 | 9.21E-08 | Crude | Mortality |
| Ratio cer18/cer24:0 | 1.711921 | 1.38603 | 2.114436 | 6.04E-07 | Crude | Mortality |
| Ratio cer20/cer24:0 | 1.582463 | 1.251101 | 2.001588 | 0.000129 | Crude | Mortality |
| Ratio cer22/cer24:0 | 1.021842 | 0.789188 | 1.323083 | 0.869799 | Crude | Mortality |
| Ratio cer24:1/cer24:0 | 1.646365 | 1.314065 | 2.062696 | 1.46E-05 | Crude | Mortality |
| Cer16 | 0.808551 | 0.587129 | 1.113476 | 0.193048 | Level 2 adjusted | Mortality |
| Cer18 | 0.711309 | 0.50909 | 0.993851 | 0.045922 | Level 2 adjusted | Mortality |
| Cer20 | 0.554122 | 0.378723 | 0.810752 | 0.002363 | Level 2 adjusted | Mortality |
| Cer22 | 0.376033 | 0.237049 | 0.596505 | 3.26E-05 | Level 2 adjusted | Mortality |
| Cer24:0 | 0.472533 | 0.306928 | 0.72749 | 0.000661 | Level 2 adjusted | Mortality |
| Cer24:1 | 0.479672 | 0.309682 | 0.742971 | 0.000999 | Level 2 adjusted | Mortality |

| | | | | | | |
|---|---|---|---|---|---|---|
| Ratio cer16/cer24:0 | 1.685295 | 1.271874 | 2.233098 | 0.000278 | Level 2 adjusted | Mortality |
| Ratio cer18/cer24:0 | 1.475002 | 1.124227 | 1.935224 | 0.00503 | Level 2 adjusted | Mortality |
| Ratio cer20/cer24:0 | 1.305177 | 0.991947 | 1.717317 | 0.057143 | Level 2 adjusted | Mortality |
| Ratio cer22/cer24:0 | 0.899397 | 0.668757 | 1.209578 | 0.483074 | Level 2 adjusted | Mortality |
| Ratio cer24:1/cer24:0 | 1.213269 | 0.934923 | 1.574483 | 0.145977 | Level 2 adjusted | Mortality |
| Cer16 | 0.876439 | 0.638934 | 1.202228 | 0.413438 | Level 1 adjusted | Mortality |
| Cer18 | 0.745996 | 0.533457 | 1.043216 | 0.08677 | Level 1 adjusted | Mortality |
| Cer20 | 0.570401 | 0.389795 | 0.834687 | 0.00385 | Level 1 adjusted | Mortality |
| Cer22 | 0.38233 | 0.242277 | 0.603343 | 3.62E-05 | Level 1 adjusted | Mortality |
| Cer24:0 | 0.459044 | 0.299389 | 0.703839 | 0.000356 | Level 1 adjusted | Mortality |
| Cer24:1 | 0.521239 | 0.343812 | 0.790228 | 0.002149 | Level 1 adjusted | Mortality |
| Ratio cer16/cer24:0 | 1.761235 | 1.343127 | 2.309497 | 4.25E-05 | Level 1 adjusted | Mortality |
| Ratio cer18/cer24:0 | 1.575472 | 1.211196 | 2.049306 | 0.000704 | Level 1 adjusted | Mortality |
| Ratio cer20/cer24:0 | 1.41128 | 1.081113 | 1.842279 | 0.011292 | Level 1 adjusted | Mortality |
| Ratio cer22/cer24:0 | 0.98258 | 0.737173 | 1.309682 | 0.90459 | Level 1 adjusted | Mortality |
| Ratio cer24:1/cer24:0 | 1.329434 | 1.027958 | 1.719325 | 0.029998 | Level 1 adjusted | Mortality |

Supplementary table 3: Hazard ratios of clinical variables for outcomes of cardiovascular events, kidney disease and all-cause mortality

| Name | Hazard Ratio | CI lower | CI upper | p-value | Model | Outcome |
|---|---|---|---|---|---|---|
| Age | 1.04859735 | 1.029608096 | 1.067936827 | 3.59477E-07 | Crude | CVE |
| BMI | 1.006075948 | 0.978871683 | 1.03403626 | 0.664932846 | Crude | CVE |
| LDL | 1.278557614 | 0.984571381 | 1.660326111 | 0.065281371 | Crude | CVE |
| Triglyceride | 1.523999724 | 1.229507624 | 1.889028676 | 0.000120113 | Crude | CVE |
| Systolic Blood Pressure | 1.017649714 | 1.006809268 | 1.02860688 | 0.001365209 | Crude | CVE |
| eGFR | 0.97776446 | 0.970628714 | 0.984952665 | 1.77758E-09 | Crude | CVE |
| $HbA_{1C}$ | 1.019498558 | 1.005663876 | 1.03352356 | 0.00560292 | Crude | CVE |
| log(UAER) | 1.860950693 | 1.454108359 | 2.381622705 | 8.03652E-07 | Crude | CVE |
| Women | 0.63585107 | 0.414516266 | 0.975369645 | 0.038060102 | Crude | CVE |
| Previous CVD | 6.332007038 | 4.20259058 | 9.54038048 | 1.09857E-18 | Crude | CVE |
| Smoking | 1.110609333 | 0.68355942 | 1.804456285 | 0.671822957 | Crude | CVE |
| Statin use | 3.051592557 | 1.825387453 | 5.101501667 | 2.08798E-05 | Crude | CVE |
| Age | 1.027873183 | 1.004334274 | 1.051963781 | 0.020026041 | Level 2 adjusted | CVE |
| BMI | 0.976684579 | 0.922859045 | 1.033649474 | 0.414684291 | Level 2 adjusted | CVE |
| LDL | 1.588591674 | 1.175023581 | 2.147721584 | 0.002627617 | Level 2 adjusted | CVE |
| Triglyceride | 0.907299246 | 0.625458957 | 1.316140592 | 0.608248557 | Level 2 adjusted | CVE |
| Systolic Blood Pressure | 1.002925346 | 0.99066586 | 1.015336544 | 0.641573735 | Level 2 adjusted | CVE |
| eGFR | 0.987074691 | 0.977809367 | 0.99642781 | 0.006857788 | Level 2 adjusted | CVE |
| $HbA_{1C}$ | 1.014804443 | 0.996974682 | 1.032953068 | 0.104174428 | Level 2 adjusted | CVE |
| log(UAER) | 1.300686918 | 0.925880067 | 1.827219873 | 0.129544061 | Level 2 adjusted | CVE |
| Women | 0.830158794 | 0.521494556 | 1.321516429 | 0.432625652 | Level 2 adjusted | CVE |
| Previous CVD | 3.720613328 | 2.33954484 | 5.916947303 | 2.84436E-08 | Level 2 adjusted | CVE |
| Smoking | 1.243178934 | 0.738039251 | 2.094053751 | 0.413248738 | Level 2 adjusted | CVE |
| Statin use | 1.481913521 | 0.820988244 | 2.674907588 | 0.191770039 | Level 2 adjusted | CVE |
| Age | 0.994790894 | 0.963785173 | 1.026794094 | 0.746484138 | Crude | Kidney failure |
| BMI | 1.015612806 | 0.978094577 | 1.054570177 | 0.419853911 | Crude | Kidney failure |
| LDL | 1.075314168 | 0.621876245 | 1.859374063 | 0.794954518 | Crude | Kidney failure |
| Triglyceride | 1.573318867 | 1.09237351 | 2.266012709 | 0.014907712 | Crude | Kidney failure |

| | | | | | | |
|---|---|---|---|---|---|---|
| Systolic Blood Pressure | 1.023421552 | 1.00199515 | 1.04530613 | 0.031985068 | Crude | Kidney failure |
| eGFR | 0.900634015 | 0.873829843 | 0.928260388 | 1.12799E-11 | Crude | Kidney failure |
| $HbA_{1C}$ | 1.034034407 | 1.010054905 | 1.058583201 | 0.005178978 | Crude | Kidney failure |
| log(UAER) | 7.09636556 | 4.191977883 | 12.01304148 | 2.9635E-13 | Crude | Kidney failure |
| Women | 0.430742011 | 0.169824396 | 1.092532546 | 0.076128718 | Crude | Kidney failure |
| Previous CVD | 2.57855195 | 1.115852933 | 5.958607953 | 0.026659517 | Crude | Kidney failure |
| Smoking | 1.695927574 | 0.697654656 | 4.122627593 | 0.243796906 | Crude | Kidney failure |
| Statin use | 2.481976223 | 0.92144758 | 6.685356935 | 0.072154476 | Crude | Kidney failure |
| Age | 0.976799843 | 0.927417368 | 1.0288118 | 0.375168078 | Level 2 adjusted | Kidney failure |
| BMI | 0.985555332 | 0.888768352 | 1.092882424 | 0.782639107 | Level 2 adjusted | Kidney failure |
| LDL | 1.22180621 | 0.746062926 | 2.000917566 | 0.426040604 | Level 2 adjusted | Kidney failure |
| Triglyceride | 1.63263548 | 0.884061816 | 3.015059084 | 0.117293711 | Level 2 adjusted | Kidney failure |
| Systolic Blood Pressure | 1.003624384 | 0.976362083 | 1.031647912 | 0.79681043 | Level 2 adjusted | Kidney failure |
| eGFR | 0.889744321 | 0.852931709 | 0.928145769 | 6.00347E-08 | Level 2 adjusted | Kidney failure |
| $HbA_{1C}$ | 1.06657952 | 1.022090166 | 1.113005399 | 0.003026142 | Level 2 adjusted | Kidney failure |
| log(UAER) | 2.21200618 | 0.95008076 | 5.15005834 | 0.065591906 | Level 2 adjusted | Kidney failure |
| Women | 0.095359811 | 0.021528727 | 0.422388813 | 0.001968508 | Level 2 adjusted | Kidney failure |
| Previous CVD | 0.922956818 | 0.301070918 | 2.829397448 | 0.888446476 | Level 2 adjusted | Kidney failure |
| Smoking | 4.360800818 | 1.03585792 | 18.35829356 | 0.044643195 | Level 2 adjusted | Kidney failure |
| Statin use | 0.53035341 | 0.146708476 | 1.917235776 | 0.333410196 | Level 2 adjusted | Kidney failure |
| Age | 1.083998219 | 1.056890322 | 1.111801399 | 4.31716E-10 | Crude | Mortality |
| BMI | 0.9619329 | 0.897492331 | 1.031000345 | 0.272633646 | Crude | Mortality |
| LDL | 0.591811567 | 0.399891195 | 0.875840567 | 0.008720662 | Crude | Mortality |
| Triglyceride | 1.194374617 | 0.851639131 | 1.675041311 | 0.303325793 | Crude | Mortality |
| Systolic Blood Pressure | 1.018228667 | 1.004384705 | 1.032263446 | 0.009699359 | Crude | Mortality |
| eGFR | 0.976337561 | 0.967228936 | 0.985531963 | 5.51755E-07 | Crude | Mortality |
| $HbA_{1C}$ | 1.006333494 | 0.986885614 | 1.026164621 | 0.526014587 | Crude | Mortality |
| log(UAER) | 1.778023368 | 1.297017954 | 2.437411978 | 0.000349019 | Crude | Mortality |
| Women | 0.718176586 | 0.419264759 | 1.230195473 | 0.228002839 | Crude | Mortality |

| | | | | | | |
|---|---|---|---|---|---|---|
| Previous CVD | 4.348374352 | 2.59778238 | 7.278654152 | 2.24282E-08 | Crude | Mortality |
| Smoking | 1.471741925 | 0.827414438 | 2.617822694 | 0.188441049 | Crude | Mortality |
| Statin use | 2.164229344 | 1.185818001 | 3.949922037 | 0.011896783 | Crude | Mortality |
| Age | 1.074534255 | 1.042945792 | 1.107079462 | 2.33499E-06 | Level 2 adjusted | Mortality |
| BMI | 0.944839947 | 0.872199777 | 1.023529871 | 0.164483548 | Level 2 adjusted | Mortality |
| LDL | 0.773872102 | 0.49395678 | 1.212409779 | 0.263093126 | Level 2 adjusted | Mortality |
| Triglyceride | 0.945882525 | 0.543626898 | 1.645786393 | 0.843916781 | Level 2 adjusted | Mortality |
| Systolic Blood Pressure | 1.002909083 | 0.987618355 | 1.018436549 | 0.710954952 | Level 2 adjusted | Mortality |
| eGFR | 0.98619113 | 0.974137915 | 0.998393482 | 0.026676461 | Level 2 adjusted | Mortality |
| $HbA_{1C}$ | 1.01200322 | 0.987318229 | 1.037305387 | 0.343638828 | Level 2 adjusted | Mortality |
| log(UAER) | 1.572327237 | 1.002034787 | 2.467192728 | 0.048975242 | Level 2 adjusted | Mortality |
| Women | 0.883934784 | 0.486845178 | 1.604905908 | 0.685172356 | Level 2 adjusted | Mortality |
| Previous CVD | 1.923971962 | 1.065709439 | 3.473430914 | 0.029923014 | Level 2 adjusted | Mortality |
| Smoking | 1.58820234 | 0.831205825 | 3.034611398 | 0.161415326 | Level 2 adjusted | Mortality |
| Statins | 0.985614826 | 0.469225082 | 2.070299786 | 0.969476886 | Level 2 adjusted | Mortality |

Supplementary table 4: Hazard ratios for ceramide and ratios for cardiovascular events separated by albuminuria status, no adjustments

| Name | Hazard Ratio | CI lower | CI upper | p-value | Model |
|---|---|---|---|---|---|
| Cer16 | 1.017821 | 0.649109 | 1.59597 | 0.938651 | Normoalbuminuria |
| Cer18 | 1.271883 | 0.825329 | 1.960051 | 0.275739 | Normoalbuminuria |
| Cer20 | 0.892414 | 0.554792 | 1.435498 | 0.63883 | Normoalbuminuria |
| Cer22 | 0.87187 | 0.538694 | 1.411112 | 0.576749 | Normoalbuminuria |
| Cer24:0 | 0.756754 | 0.456721 | 1.253887 | 0.279337 | Normoalbuminuria |
| Cer24:1 | 0.949629 | 0.591326 | 1.525038 | 0.830669 | Normoalbuminuria |
| Ratio Cer16/Cer24:0 | 1.438587 | 0.948065 | 2.1829 | 0.087397 | Normoalbuminuria |
| Ratio Cer18/Cer24:0 | 1.781448 | 1.228911 | 2.582415 | 0.002303 | Normoalbuminuria |
| Ratio Cer20/Cer24:0 | 1.503093 | 0.978989 | 2.307779 | 0.062477 | Normoalbuminuria |
| Ratio Cer22/Cer24:0 | 1.60787 | 1.03071 | 2.508218 | 0.036323 | Normoalbuminuria |
| Ratio Cer24:1/Cer24:0 | 1.719364 | 1.116846 | 2.646928 | 0.013817 | Normoalbuminuria |
| Cer16 | 1.541817 | 1.076474 | 2.208322 | 0.018178 | Moderately Increased |
| Cer18 | 1.803756 | 1.310159 | 2.483314 | 0.000299 | Moderately Increased |
| Cer20 | 1.303163 | 0.922516 | 1.840872 | 0.133 | Moderately Increased |
| Cer22 | 1.258576 | 0.887204 | 1.785397 | 0.197357 | Moderately Increased |
| Cer24:0 | 1.300178 | 0.92795 | 1.821718 | 0.127153 | Moderately Increased |
| Cer24:1 | 1.539388 | 1.141207 | 2.076501 | 0.004729 | Moderately Increased |
| Ratio Cer16/Cer24:0 | 1.060735 | 0.760474 | 1.47955 | 0.728386 | Moderately Increased |
| Ratio Cer18/Cer24:0 | 1.195981 | 0.875033 | 1.634649 | 0.261607 | Moderately Increased |
| Ratio Cer20/Cer24:0 | 1.008452 | 0.716747 | 1.418877 | 0.961469 | Moderately Increased |
| Ratio Cer22/Cer24:0 | 0.891984 | 0.629855 | 1.263203 | 0.519663 | Moderately Increased |
| Ratio Cer24:1/Cer24:0 | 1.245724 | 0.925808 | 1.676189 | 0.146806 | Moderately Increased |
| Cer16 | 1.295086 | 0.995982 | 1.684014 | 0.053618 | Severely Increased |
| Cer18 | 1.10577 | 0.851478 | 1.436004 | 0.450802 | Severely Increased |
| Cer20 | 1.034866 | 0.780562 | 1.372023 | 0.811735 | Severely Increased |
| Cer22 | 1.071787 | 0.808881 | 1.420143 | 0.629227 | Severely Increased |
| Cer24:0 | 1.110295 | 0.832182 | 1.481354 | 0.476955 | Severely Increased |
| Cer24:1 | 1.089437 | 0.82349 | 1.441273 | 0.548569 | Severely Increased |
| Ratio Cer16/Cer24:0 | 1.022587 | 0.766645 | 1.363975 | 0.879212 | Severely Increased |
| Ratio Cer18/Cer24:0 | 0.956326 | 0.709624 | 1.288796 | 0.769257 | Severely Increased |
| Ratio Cer20/Cer24:0 | 0.897105 | 0.669335 | 1.202384 | 0.467461 | Severely Increased |
| Ratio Cer22/Cer24:0 | 0.907777 | 0.681311 | 1.20952 | 0.508733 | Severely Increased |
| Ratio Cer24:1/Cer24:0 | 0.970632 | 0.715171 | 1.317343 | 0.848302 | Severely Increased |
